# Cognitive pleiotropy reveals disorder-specific and shared biology for schizophrenia and bipolar disorder

**DOI:** 10.64898/2026.07.07.26357446

**Authors:** Upasana Bhattacharyya, Jibin John, Michael Preuss, Max Lam, Todd Lencz

**Author notes:** Corresponding Author: Todd Lencz; Office: Division of Psychiatry Research, Zucker Hillside Hospital, Glen Oaks, NY, USA 11004. These authors contributed equally (Upasana Bhattacharyya, Jibin John).

## Abstract

Schizophrenia (SCZ) and bipolar disorder (BIP) share substantial common-variant liability but differ in clinical course, cognition, and treatment response. We previously resolved this overlap into three cognitively divergent components, SCZcondBIP (negatively correlated with cognition and education), BIPcondSCZ (positively correlated with both), and PSY-shared (negative with cognition, positive with education), representing distinct neurocognitive axes. Here, we applied directional pleiotropic meta-analysis (PLEIO) to integrate these components with cognitive task performance and educational attainment, yielding 818 consensus loci of which 514 (63%) were shared across all three components and 220 were component-specific, including 99 novel loci absent from individual GWAS. Each component was partitioned into concordant (aligning with the expected cognitive direction, e.g., increased cognition with reduced SCZcondBIP risk) and discordant (reverse pattern, e.g., increased cognition with increased SCZcondBIP risk) locus sets. Pathway analyses revealed marked biological divergence: SCZcondBIP concordant loci were enriched for neurodevelopmental pathways and discordant loci for cellular-homeostasis pathways, suggesting two separable processes: an early developmental-regulatory branch linking cognitive disadvantage to increased risk, and a homeostatic/metabolic-stress branch enabling cognitive advantage despite increased risk. BIPcondSCZ concordant loci, where increased cognition co-occurs with increased bipolar risk, were enriched for synaptic pathways. Unlike neurodevelopmental pathways, which emerged as primary cognitive determinants in SCZcondBIP, synaptic pathways may principally mediate disease liability without substantially impacting cognition, explaining preserved or enhanced performance alongside increased bipolar risk. Discordant loci, where decreased cognition co-occurs with decreased bipolar risk; implicated cellular-homeostasis pathways through mitochondria mediated pathways, consistent with mitochondrial dysfunction and reduced cellular resilience contributing to bipolar pathophysiology and progressive cognitive decline. PSY-shared concordant loci showed nominal synaptic and cellular-homeostasis enrichment, while discordant loci implicated neuroimmune and vesicular processes, directionally consistent with disorder-specific partitions. Schizophrenia and bipolar disorder genetic risk comprises biologically coherent disorder-specific and shared dimensions; integrating these with cognition exposes directional pleiotropic architecture across divergent developmental, synaptic, and metabolic pathways, providing a mechanistic framework for understanding cognitive heterogeneity in severe psychiatric illness.

## Introduction

Schizophrenia (SCZ) and bipolar disorder (BIP) are severe psychiatric disorders that impose a substantial global burden worldwide^1–3^, typically emerging during late adolescence or early adulthood—a critical period for educational and social development^4,5^. Both disorders are highly heritable, with twin-study estimates ranging from 60–80% for SCZ^6,7^ and 60–85% for BIP^8,9^. Although the two disorders are strongly genetically correlated (rg ∼ 0.67)^10^, reflecting an extensive shared biology^11–14^, they retain highly distinct symptom profiles, longitudinal trajectories, and treatment responses^15–19^. This clinical heterogeneity, combined with their incomplete genetic overlap, underscores that a shared transdiagnostic liability cannot fully account for the unique pathophysiology of either disorder^20^.

To disentangle shared and disorder-specific genetic risk, we previously applied bidirectional multi-trait conditional and joint analysis (mtCOJO)^21^ to the largest available GWAS for SCZ^22^ and BIP^8^, conditioning each disorder on the other to recover independent genetic signals (SCZcondBIP, BIPcondSCZ)(MEDRXIV/2026/357436), and used Genomic Structural Equation Modeling (Genomic SEM)^23^ to derive a shared latent psychosis factor (PSY-shared)(MEDRXIV/2026/357436). Clinically, the two disorders diverge sharply in cognitive trajectory: schizophrenia is associated with marked premorbid cognitive deficits often detectable in childhood^24^, whereas individuals who later develop bipolar disorder typically exhibit average or above-average premorbid functioning, with impairments emerging primarily after illness onset^24,25^. Yet this clinical divergence is not mirrored in genetic correlation analyses, where both disorders show a familiar paradox: a negative genetic correlation with cognition alongside a positive correlation with education, leaving open whether this pattern reflects a single shared mechanism or a composite of distinct dimensions. Partitioning resolved this ambiguity.

SCZcondBIP was negatively correlated with both cognition and education, consistent with schizophrenia-specific liability being tied to early neurocognitive burden and reduced premorbid cognitive function. BIPcondSCZ, by contrast, was positively correlated with both, suggesting that once shared psychosis liability is removed, bipolar-specific risk is less coupled to cognitive compromise and more compatible with preserved premorbid cognitive functioning. The PSY-shared factor retained the mixed profile (negative with cognition, positive with education), implying that shared liability drives the paradox observed in cross-disorder analyses. Together, these results indicate that cognitive outcome depends on the relative balance of schizophrenia-specific, bipolar-specific, and shared psychosis influences rather than on overall liability alone, and that cognitive performance and educational attainment index partly distinct genetic influences whose joint leverage may help disentangle pleiotropic pathways underlying psychiatric risk.

Independently, a Genomic SEM–based GWAS-by-subtraction approach partitioned schizophrenia and bipolar liability into a SCZ-specific component and a component shared with BIP^26^, revealing similarly opposing genetic relationships with educational attainment: SCZ-specific risk was negatively correlated, whereas the shared SCZ–BIP component was positively correlated. These convergent findings suggest that the previously observed positive genetic correlation between overall SCZ liability and educational attainment^27^ is largely driven by genetic factors shared with BIP rather than by SCZ-specific risk. However, that framework did not explicitly model BIP-specific liability, leaving its unique genetic architecture less well characterized.

In prior work, we leveraged the pleiotropic relationship between SCZ, cognitive task performance, and educational attainment to partition SCZ risk loci into “concordant” variants (where risk alleles associate with the expected reduction in cognitive and educational performance) and “discordant” variants (where risk alleles paradoxically associate with higher performance). This directional dissection revealed a biological dichotomy: concordant loci preferentially implicate prenatal neurodevelopmental pathways, whereas discordant loci are enriched for postnatally expressed synaptic mechanisms^28,29^. Motivated by these converging lines of evidence, the present study extends our pleiotropic framework by applying directional pleiotropic meta-analysis (PLEIO) to integrate intermediate cognitive phenotypes (cognitive task performance and educational attainment) with SCZcondBIP, BIPcondSCZ, and PSY-shared. We then characterize the implicated biology through downstream gene-set and drug-enrichment analyses, aiming to delineate the divergent developmental architectures underlying cognitive heterogeneity across these severe psychiatric disorders.

## Methods

### Overview of Methodological Framework

We previously applied bidirectional multi-trait conditional and joint analysis (mtCOJO)^21^ to the largest available GWAS for SCZ^22^ and BIP^8^, conditioning each disorder on the other to recover independent genetic signals (SCZcondBIP, BIPcondSCZ)(MEDRXIV/2026/357436), and used Genomic Structural Equation Modeling (Genomic SEM)^23^ to derive a shared latent psychosis factor (PSY-shared)(MEDRXIV/2026/357436). The three components diverged sharply in their cognitive profiles: SCZcondBIP was negatively correlated with both cognition and education, BIPcondSCZ was positively correlated with both, and PSY-shared showed a mixed pattern, negative with cognition but positive with education(MEDRXIV/2026/357436). This contrasts with the unpartitioned disorders, which both show the same paradoxical pattern of negative correlation with cognition alongside positive correlation with education, indicating that the decomposition resolves this shared paradox into distinct neurocognitive axes that are masked at the disorder level(MEDRXIV/2026/357436). Thus, in this study, to leverage these divergent cognitive profiles for enhanced locus discovery and to resolve the directional heterogeneity underlying cognitive outcomes across the two disorders, we applied the directionally informative pleiotropic meta-analytic tool, Pleiotropic Locus Exploration and Interpretation using Optimal test (PLEIO)^30^, integrating each of the three components with cognitive task performance (CTP)^31^ and educational attainment (EDU)^27^.Pleiotropic loci obtained from each meta-analysis were partitioned into concordant and discordant sets, defined relative to the sign of each component’s genetic correlation with CTP and EDU. For SCZcondBIP, which is negatively correlated with both CTP and EDU, concordant loci are those where alleles that increase CTP and EDU also decrease SCZcondBIP liability (i.e., the expected direction), whereas discordant loci are those where alleles that increase CTP and EDU paradoxically increase SCZcondBIP liability. For BIPcondSCZ, which is positively correlated with both CTP and EDU, concordant loci are those where alleles that increase CTP and EDU also increase BIPcondSCZ liability, whereas discordant loci are those where alleles that decrease CTP and EDU also decrease BIPcondSCZ liability, opposing the expected positive relationship. For PSY-shared, which shows a mixed correlation profile (negative with CTP, positive with EDU), concordant loci are those where alleles that decrease CTP and/or increase EDU also increase PSY-shared liability, whereas discordant loci are those where alleles that increase CTP and/or decrease EDU also increase PSY-shared liability. Finally, we functionally characterized these groups through genomic risk-locus definition and candidate-gene mapping (FUMA, version 2.1.5)^32^, identification of novel loci against published GWAS (BEDTools)^33^, competitive gene-set enrichment (MAGMA)^34^, and drug-target and druggability assessment (DGIdb version 5.0/Enrichr)^35–38^ (Figure 1). The analytical workflow is detailed in the Supplementary Methods section.

**Figure 1.**
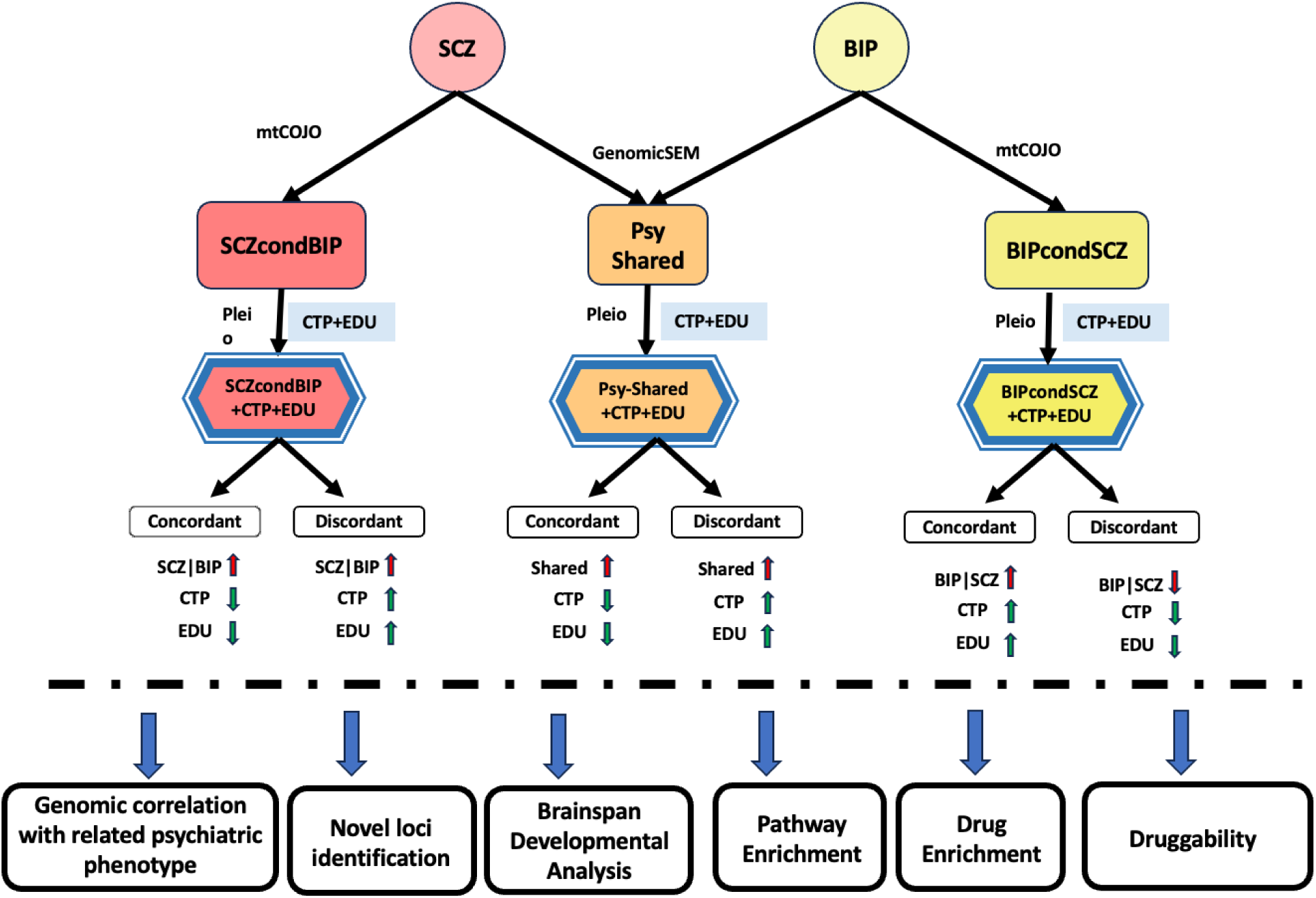
Study design overview. Summary GWAS statistics for schizophrenia (Trubetskoy et al., 2022) and bipolar disorder (O’Connell/Koromina et al., 2025) were decomposed into three genetically distinguishable liability components: a schizophrenia-conditional component (SCZcondBIP), derived by conditioning schizophrenia on bipolar disorder using mtCOJO; a bipolar-conditional component (BIPcondSCZ), derived by conditioning bipolar disorder on schizophrenia using mtCOJO; and a shared psychosis component (PSY-shared), derived by modelling the schizophrenia–bipolar disorder genetic covariance in Genomic SEM. Each component was then jointly meta-analysed with cognitive task performance (CTP) and educational attainment (EDU) using PLEIO, and the resulting pleiotropic SNPs were partitioned into concordant (effect direction on the component aligns with the expected direction on CTP/EDU) and discordant (opposite direction) sets, yielding six directional partitions in addition to three unpartitioned component-level sets. Downstream analyses—genetic correlations with related psychiatric phenotypes, novel-locus identification, BrainSpan developmental-timing analysis, MAGMA pathway enrichment, Enrichr drug-target enrichment, and DGIdb druggability annotation—were applied to each of the resulting nine gene sets.

## Result

### Genomic risk loci and cross-component overlap

#### Stage 1: PLEIO-informed loci

PLEIO meta-analysis of each component with cognitive task performance (CTP) and educational attainment (EDU) substantially expanded the set of implicated loci for each component. SCZcondBIP was mapped to 635 loci, BIPcondSCZ to 597 loci, and the PSY-shared factor to 734 loci (Figure 2a; Supplementary Table 1a). Merging these PLEIO-derived loci across components yielded 818 distinct consensus loci, of which 514 (63%) were shared across all three components. An additional 84 loci were shared by exactly two components (38 between SCZcondBIP and the PSY-shared factor, 27 between the two conditional components, and 19 between BIPcondSCZ and the PSY-shared factor). The remaining 220 loci were component-specific: 157 unique to the PSY-shared factor, 43 to SCZcondBIP, and 20 to BIPcondSCZ (Figure 2b; Supplementary Table 1a).

**Figure 2.**
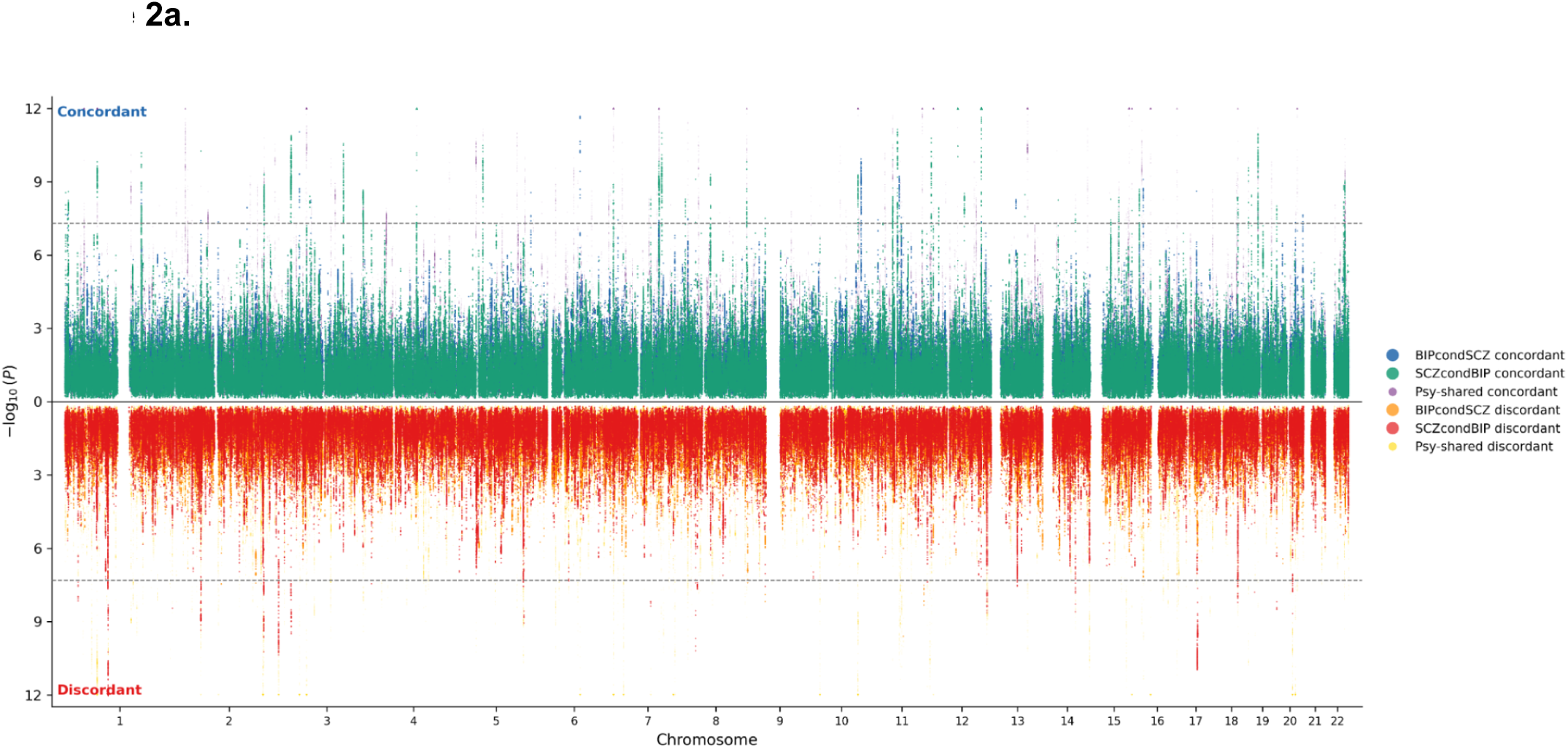
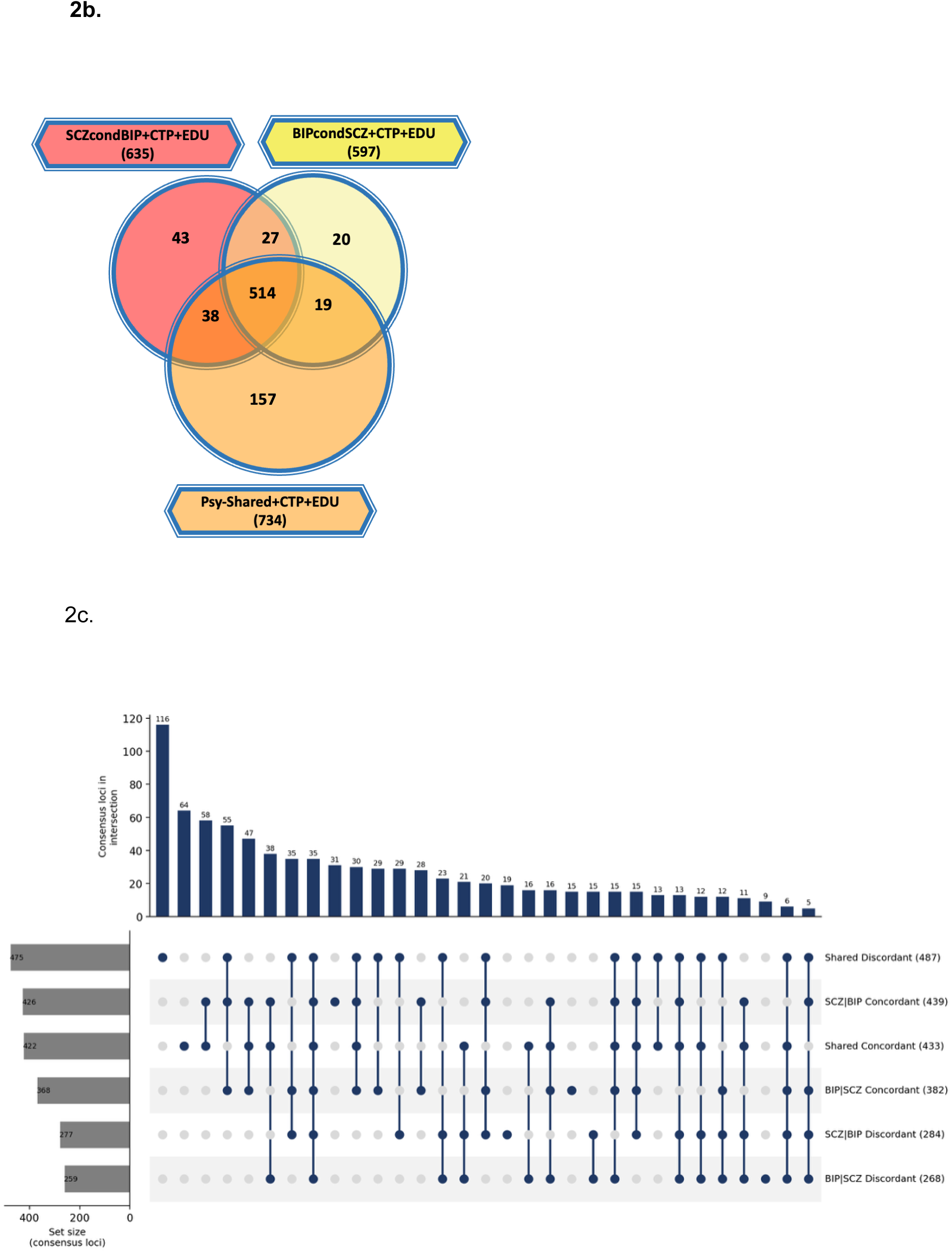
Pleiotropic locus landscape across components and partitions. (a) Miami plot of −log₁₀(P) values from PLEIO meta-analysis of each component with CTP and EDU. The upper half displays concordant variants and the lower half discordant variants; colours distinguish the six partitions (SCZcondBIP concordant/discordant, BIPcondSCZ concordant/discordant, PSY-shared concordant/discordant). (b) Venn diagram showing overlap of independent loci identified after separate meta-analysis of each component (SCZcondBIP, BIPcondSCZ, PSY-shared) with CTP and EDU, alongside loci from the unpartitioned schizophrenia and bipolar disorder GWAS. (c) UpSet plot quantifying the intersections among the six concordant/discordant partitions (SCZcondBIP concordant, SCZcondBIP discordant, BIPcondSCZ concordant, BIPcondSCZ discordant, PSY-shared concordant, PSY-shared discordant).

#### Stage 2: Directionally partitioned locus sets

Following pleiotropic meta-analysis, we partitioned each component into concordant and discordant locus sets based on the direction of genetic correlations with CTP and EDU. For the schizophrenia-conditional component (SCZcondBIP), which was negatively correlated with both CTP and EDU, concordant variants were defined as those that increase CTP and EDU while reducing schizophrenia-specific liability, whereas discordant variants were defined as those that increase CTP and EDU but paradoxically also increase schizophrenia-specific liability. For the bipolar-conditional component (BIPcondSCZ), which was positively correlated with both CTP and EDU, concordant variants were defined as those that increase CTP and EDU and also increase bipolar-specific liability, whereas discordant variants were defined as those that decrease CTP and EDU and also decrease bipolar-specific liability. The PSY-shared factor resembled unpartitioned schizophrenia, being negatively correlated with CTP but positively correlated with EDU. Given this mixed profile, and consistent with our previous work, concordant variants for the shared factor were defined as those in which alleles decrease CTP and/or increase EDU while also increasing PSY-shared liability, whereas discordant variants were those in which alleles increase CTP and/or decrease EDU while also increasing PSY-shared liability.

This partitioning yielded six locus sets: for SCZcondBIP, 439 concordant and 284 discordant loci; for BIPcondSCZ, 382 concordant and 268 discordant loci; and for the PSY-shared factor, 433 concordant and 487 discordant loci. Merging these six sets at a minimum overlap of one base pair yielded 867 consensus loci (Figure 2c; Supplementary Table 1b). Of these, 254 (29.3%) were specific to a single directional set and 613 (70.7%) were shared across two or more sets. Among set-specific loci, 116 were unique to the shared discordant set and 64 to the shared concordant set, followed by 31 unique to SCZcondBIP concordant, 19 to SCZcondBIP discordant, 15 to BIPcondSCZ concordant, and 9 to BIPcondSCZ discordant loci (Figure 2c; Supplementary Table 1b).

The overlap structure followed the expected directional logic, with same-direction sets converging across components. Among concordant sets, 58 consensus loci were shared exclusively between SCZcondBIP concordant and shared concordant loci, 28 exclusively between SCZcondBIP concordant and BIPcondSCZ concordant loci, and 47 were common to all three concordant sets. Among discordant sets, 29 consensus loci were shared exclusively between SCZcondBIP discordant and shared discordant loci, 15 exclusively between SCZcondBIP and BIPcondSCZ discordant loci, and 23 were common to all three discordant sets. A core of 35 consensus loci was common to all six sets (Figure 2c; Supplementary Table 1b).

In contrast, concordant and discordant sets within the same component rarely coincided, with only 1 consensus locus overlapping between SCZcondBIP concordant and SCZcondBIP discordant loci, and 13 between shared concordant and shared discordant loci. However, a number of consensus loci spanned sets with opposite directional labels across components: for example, 29 loci were shared between BIPcondSCZ concordant and shared discordant sets, and 16 between BIPcondSCZ discordant and shared concordant sets (Figure 2c; Supplementary Table 1b).

### Identification of novel loci

PLEIO meta-analysis of each component with cognitive task performance and educational attainment identified 145 novel pleiotropic loci (80 for the PSY-shared component, 34 for the schizophrenia-conditional component, and 31 for the bipolar-conditional component) that did not overlap any genome-wide significant locus from the schizophrenia (PGC3), bipolar disorder (PGC-BIP), cognitive task performance, or EA3 GWAS and did not lie within 20 kb of any EA4 lead SNP. Merging these novel loci across components yielded 99 distinct novel genomic regions, 17 of which were shared as novel across all three components. A further 312 loci (122 for the PSY-shared component, 110 for the schizophrenia-conditional component, and 80 for the bipolar-conditional component) overlapped none of the four reference GWAS but did lie within 20 kb of an EA4 lead SNP, and are therefore reported as EA4-adjacent rather than confirmed novel; these corresponded to 175 distinct genomic regions (Supplementary Table 2).

### Gene-set analysis (MAGMA)

To investigate the shared and distinct biological pathophysiologies underlying each component and its directional partitions, we performed competitive gene-set analysis against Gene Ontology (GO) gene sets. Enriched gene sets were grouped into three biological categories—synaptic structure and signalling, neurodevelopmental processes (including transcriptional, epigenetic, and RNA-processing regulation), and cellular homeostasis and metabolic support—and each locus set was characterized by the category in which it was predominantly enriched.

Within the schizophrenia-conditional component, the concordant loci (26 gene sets at FDR < 0.05) were enriched overwhelmingly in neurodevelopmental processes, including sequence-specific DNA binding (FDR-P < 0.001), positive regulation of RNA metabolic process (FDR-P = 0.001), GABAergic neuron differentiation (FDR-P = 0.005), beta-catenin binding (FDR-P = 0.005), diencephalon development (FDR-P = 0.006), forebrain neuron differentiation (FDR-P = 0.016), and forebrain neuron fate commitment (FDR-P = 0.017), with the remaining sets distributed across synaptic structure. The discordant loci were enriched in cellular homeostasis and metabolic support, with two gene sets significant at FDR < 0.05 (nuclear protein-containing complex, FDR-P = 0.007; response to oxygen–glucose deprivation, FDR-P = 0.018) and leading nominal sets including UMP catabolic process (P = 2.32 × 10⁻⁵) and dUMP metabolic process (P = 2.88 × 10⁻⁵) (Figure 3; Supplementary Table 3).

**Figure 3.**
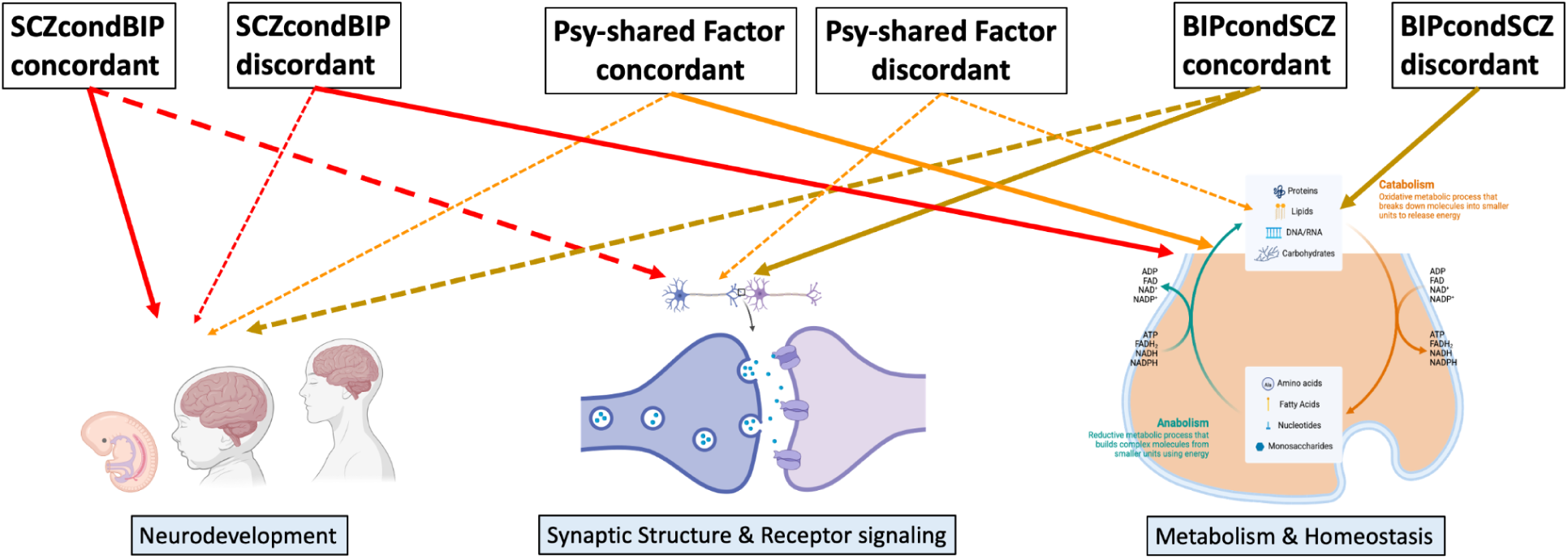
Biological pathway enrichment across concordant and discordant partitions. (a) MAGMA gene-set enrichment results across the six concordant/discordant partitions. Arrows and bars link each partition to broad pathway categories: solid lines denote major enriched pathway groups, dashed lines indicate additional pathway groups with fewer enrichments, and thin dotted lines denote pathway groups that were enriched but did not reach FDR significance.

Within the bipolar-conditional component, the concordant loci yielded four gene sets at FDR < 0.05 and were enriched in synaptic structure and receptor signalling (synaptic membrane adhesion, FDR-P = 0.039; neurotrophin TRK receptor binding, FDR-P = 0.039) together with the neurodevelopmental set cranial ganglion development (FDR-P = 0.039) and fungiform papilla development (FDR-P = 0.049). The discordant loci were enriched in cellular homeostasis and metabolic support, with four gene sets at FDR < 0.05 dominated by mitochondrial and apoptotic processes: intrinsic apoptotic signalling pathway in response to hypoxia (FDR-P < 0.001), mitochondrial fragmentation involved in apoptotic process (FDR-P = 0.017), pyrimidine deoxyribonucleoside metabolic process (FDR-P = 0.027), and mitochondrial protein catabolic process (FDR-P = 0.049) (Figure 3; Supplementary Table 3).

Within the shared component, the concordant loci yielded three gene sets at FDR < 0.05 and were enriched in neurodevelopmental processes (regulation of cardiac muscle tissue development, FDR-P = 0.001; protein–DNA complex, FDR-P = 0.001), with ErbB-3 class receptor binding (FDR-P = 0.001) representing synaptic receptor signalling. The discordant loci yielded no gene set at FDR < 0.05; their leading nominal associations were enriched in neuro-immune interactions (B-1 B cell differentiation, P = 8.49 × 10⁻⁶; B cell proliferation involved in immune response, P = 2.32 × 10⁻⁵) and synaptic structure (neurotransmitter loading into synaptic vesicle, P = 1.61 × 10⁻⁵) (Figure 3; Supplementary Table 3).

### Developmental age

We next examined the developmental timing of the genes implicated by each component and directional partition using MAGMA gene-property analysis across BrainSpan developmental periods. The SCZcondBIP concordant loci showed no FDR-significant enrichment at any developmental period; at nominal significance, a trend toward early prenatal enrichment was observed (P = 0.015, FDR-P = 0.160). The SCZcondBIP discordant loci showed no enrichment at any period at either FDR or nominal significance (all P > 0.05). The BIPcondSCZ concordant loci showed robust FDR-significant enrichment spanning the early prenatal (FDR-P = 0.003), early-mid-prenatal (FDR-P = 0.003), and late-mid-prenatal (FDR-P = 0.003) periods; no additional nominal trends were observed. The BIPcondSCZ discordant loci showed parallel FDR-significant enrichment in the early prenatal (FDR-P = 0.007), early-mid-prenatal (FDR-P = 0.013), and late-mid-prenatal (FDR-P = 0.005) periods; no additional nominal trends were observed. The PSY-shared concordant loci showed no FDR-significant enrichment at any period and no nominal trends (all P > 0.05). The PSY-shared discordant loci showed no FDR-significant enrichment; at nominal significance, trends were observed in the early prenatal (P = 0.039, FDR-P = 0.427) and late-mid-prenatal (P = 0.032, FDR-P = 0.177) periods (Supplementary Table 4).

### Drug-target enrichment analysis (DGIdb/Enrichr)

As a positive control for the biological relevance of the implicated gene sets, we tested each concordant and discordant partition for enrichment of drug targets in DGIdb via Enrichr, evaluated at nominal significance (P < 0.05) (Supplementary Table 5).

Schizophrenia-conditional concordant loci. The concordant loci of the schizophrenia-conditional component were enriched for the phenothiazine antipsychotics fluphenazine (P = 0.023), triflupromazine (P = 0.047), and promethazine (P = 0.023) and for the atypical antipsychotics clozapine (P = 0.030) and aripiprazole (P = 0.037). The repurposing layer included the anxiolytic buspirone (P = 0.025), the gabapentinoid pregabalin (P = 0.030), the stimulant methylphenidate (P = 0.032), and the L-type calcium channel blockers nimodipine (P = 0.048), nicardipine (P = 0.037), and cinnarizine (P = 0.037)—the last of mechanistic interest given the established association of L-type calcium channel genes with both disorders (Supplementary Table 5).

Schizophrenia-conditional discordant loci. The discordant loci of the schizophrenia-conditional component were enriched for the atypical antipsychotic risperidone (P = 0.002). We also found genes in this group to be enriched for drugs currently being explored in schizophrenia as adjunctive treatment or repurposing candidates, including the anticonvulsant mood stabilisers oxcarbazepine (P = 0.011), topiramate (P = 0.014), and zonisamide (P = 0.033), together with the dopaminergic anti-Parkinson agent safinamide (P = 0.041) and the anti-inflammatory antioxidant curcumin (P = 0.026) (Supplementary Table 5).

Bipolar-conditional concordant loci. The concordant loci of the bipolar-conditional component showed no antipsychotic enrichment, implicating instead the tricyclic antidepressant protriptyline (P = 0.036) and the immunomodulatory cytokine interferon beta-1b (P = 0.047); the remaining enrichment was observed for proton-pump inhibitors (omeprazole, P = 0.045; lansoprazole, P = 0.049) and oncology-related agents (afatinib, P = 0.023; dinaciclib, P = 0.036; pertuzumab, P = 0.036) (Supplementary Table 5).

Bipolar-conditional discordant loci. The discordant loci of the bipolar-conditional component were enriched for the antipsychotics fluspirilene (P = 0.012), fluphenazine (P = 0.020), prochlorperazine (P = 0.045), and risperidone (P = 0.041). This group, like the schizophrenia-conditional discordant loci, also showed enrichment for drugs suggested for repurposing, including dopaminergic and glutamatergic agents—the anti-Parkinson agents safinamide (P = 0.033), carbidopa (P = 0.035), and domperidone (P = 0.039), the glutamate modulator riluzole (P = 0.033), and the system-xc⁻ agent sulfasalazine (P = 0.019)—together with the anticonvulsant oxcarbazepine (P = 0.008) and the stimulant methylphenidate (P = 0.003) (Supplementary Table 5).

Shared concordant loci. The concordant loci of the shared component were enriched for the phenothiazine antipsychotics fluphenazine (P = 0.003), promethazine (P = 0.007), triflupromazine (P = 0.010), and prochlorperazine (P = 0.015) and for the atypical antipsychotic risperidone (P = 0.021). In addition, these genes were enriched for the L-type calcium channel blockers nimodipine (P = 0.017), nicardipine (P = 0.044), and nifedipine (P = 0.049), the selective estrogen receptor modulator raloxifene (P = 0.044), the neurosteroid progesterone (P = 0.009), the system-xc⁻ agent sulfasalazine (P = 0.008), and the gabapentinoid pregabalin (P = 0.040) (Supplementary Table 5).

Shared discordant loci. The discordant loci of the shared component were enriched for the atypical antipsychotic amisulpride (P = 0.024). These genes were also enriched for the tricyclic antidepressant amitriptyline (P = 0.033), the gabapentinoid pregabalin (P = 0.020), the L-type calcium channel blocker cinnarizine (P = 0.012), and a cluster of immune and anti-inflammatory agents—sulfasalazine (P = 0.029), celecoxib (P = 0.025), curcumin (P = 0.010), and lenalidomide (P = 0.012) (Supplementary Table 5).

### Druggability of the implicated genes

To provide a more granular assessment of therapeutic tractability, the FUMA-mapped candidate genes from each of the six concordant and discordant partitions were annotated against the Drug–Gene Interaction module of DGIdb (Supplementary Table 6a–f). Across the six partitions, both the number of FUMA-mapped genes with any recorded drug–gene interaction and the number targeted by at least one drug with current regulatory approval were substantial and comparable in magnitude: 1,298 interacting genes for SCZcondBIP concordant (966 with ≥1 approved-drug interaction; Supplementary Table 6a), 837 for SCZcondBIP discordant (630 approved-drug-targeted; Supplementary Table 6b), 1,125 for BIPcondSCZ concordant (828 approved-drug-targeted; Supplementary Table 6c), 790 for BIPcondSCZ discordant (586 approved-drug-targeted; Supplementary Table 6d), 1,370 for PSY-shared concordant (1,034 approved-drug-targeted; Supplementary Table 6e), and 1,400 for PSY-shared discordant (1,054 approved-drug-targeted; Supplementary Table 6f). Beyond the antipsychotic, mood-stabilising, and anti-inflammatory classes highlighted in the drug-target enrichment analysis above, the DGIdb annotations across the six partitions collectively spanned multiple additional neuropharmacological mechanisms represented by approved compounds, including dopaminergic agents (e.g., the dopamine agonists apomorphine, rotigotine, and cabergoline and the vesicular monoamine transporter inhibitors reserpine and tetrabenazine), cholinergic agents (e.g., the acetylcholinesterase inhibitors donepezil and galantamine and the nicotinic agents mecamylamine and varenicline), α2-adrenergic agonists (clonidine, guanfacine), monoaminergic antidepressants (mirtazapine, duloxetine, venlafaxine, and tricyclic agents), glutamatergic and neuroprotective agents (riluzole, ibudilast), L-type calcium channel blockers (nimodipine, nicardipine, isradipine), and anti-inflammatory and immunomodulatory agents (the cyclooxygenase inhibitors naproxen and ketorolac and interferon beta-1a and beta-1b) (Supplementary Table 6a–f).

## Discussion

Schizophrenia (SCZ) and bipolar disorder (BIP) share a substantial fraction of their common-variant genetic risk (rg ≈ 0.67), yet differ in symptom profile, course, treatment response, and relationship to cognition and educational attainment. Applying bidirectional mtCOJO and Genomic SEM to the largest available GWAS of SCZ and BIP, we previously resolved this joint architecture into three components: an SCZ-specific factor (SCZcondBIP), a BIP-specific factor (BIPcondSCZ), and a shared psychosis factor (PSY-shared)(MEDRXIV/2026/357436). The three components diverged in their genetic correlations with cognitive task performance (CTP) and educational attainment (EDU)(MEDRXIV/2026/357436). These three components diverge in their correlations with a range of traits, including the most important endophenotypes, cognition and educational attainment(MEDRXIV/2026/357436), which may explain heterogeneous cognitive outcomes across the two disorders. Originally, SCZ and BIP both show a paradoxical relationship with cognitive traits, a negative correlation with cognition alongside a positive correlation with education, leaving open whether this reflects one shared mechanism or a composite of distinct dimensions. After resolving the disease risk into disease-specific and shared factors, SCZcondBIP was negatively correlated with both cognition and education, consistent with the view that schizophrenia-specific liability is tied to early neurocognitive burden and impaired premorbid functioning. BIPcondSCZ, by contrast, was positively correlated with both, suggesting that once shared psychosis liability is removed, bipolar-specific risk is less coupled to early cognitive compromise and more compatible with preserved premorbid functioning. The PSY-shared factor retained the mixed profile (negative with cognition, positive with education), implying that shared liability drives the paradox seen in cross-disorder analyses. Together, these results suggest cognitive outcome depends on the relative balance of schizophrenia-specific, bipolar-specific, and shared psychosis influences rather than on overall liability alone. Motivated by this decomposition, we extended our previous framework using directional pleiotropic meta-analysis (PLEIO) to integrate each component with CTP and EDU to extend our previous work^28,29^. This approach both enhances locus discovery and lets us partition the resulting SNPs into concordant and discordant groups relative to each component’s genomic correlation with cognition and education. Tracing these partitions through functional and developmental annotation reveals divergent architectures that illuminate clinical and cognitive heterogeneity even within the same disorder. both show a paradoxical relationship with cognitive traits, a negative correlation with cognition alongside a positive correlation with education, leaving open whether this reflects one shared mechanism or a composite of distinct dimensions. After resolving the disease risk into disease-specific and shared factors, SCZcondBIP was negatively correlated with both cognition and education, consistent with the view that schizophrenia-specific liability is tied to early neurocognitive burden and impaired premorbid functioning. BIPcondSCZ, by contrast, was positively correlated with both, suggesting that once shared psychosis liability is removed, bipolar-specific risk is less coupled to early cognitive compromise and more compatible with preserved premorbid functioning. The PSY-shared factor retained the mixed profile (negative with cognition, positive with education), implying that shared liability drives the paradox seen in cross-disorder analyses. Together, these results suggest cognitive outcome depends on the relative balance of schizophrenia-specific, bipolar-specific, and shared psychosis influences rather than on overall liability alone. We thus used these cognitive traits to extend our previous framework[33, 34] by applying directional pleiotropic meta-analysis (PLEIO) to integrate each component with cognitive and educational traits. This approach both enhances locus discovery and lets us partition the resulting SNPs into concordant and discordant groups relative to each component’s genomic correlation with cognition and education. Tracing these partitions through functional and developmental annotation reveals divergent architectures that illuminate clinical and cognitive heterogeneity even within the same disorder.

Locus discovery showed that PLEIO meta-analysis of each component with cognitive task performance (CTP) and educational attainment (EDU) substantially expanded the set of implicated loci for each component. SCZcondBIP was mapped to 635 loci, BIPcondSCZ to 597 loci, and the PSY-shared factor to 734 loci. Merging these PLEIO-derived loci across components yielded 818 distinct consensus loci, of which a substantial portion (220 loci) were component-specific: 157 unique to the PSY-shared factor, 43 to SCZcondBIP, and 20 to BIPcondSCZ. Of these, 99 pleiotropic loci were novel across all three components. Of these 99 novel pleiotropic loci, 76 were later identified in the recent education GWAS^39^, which has substantially larger sample size than EA3^27^, used as input for the pleiotropic meta-analysis in this study. This pattern suggests that integrating psychiatric liability with cognition and education boosts sensitivity to loci that would otherwise require much larger samples. Two features of this locus inventory deserve emphasis. First, the strong asymmetry in component-specific loci, 157 unique to the PSY-shared factor versus 43 for SCZcondBIP and only 20 for BIPcondSCZ, likely reflects differences in statistical power across the three components. The PSY-shared factor, which aggregates signal from both disorders, is expected to have the greatest power to detect pleiotropic loci with cognition and education, yielding the largest number of unique associations. The comparatively small BIPcondSCZ-unique fraction is consistent with the moderate sample size of the bipolar disorder GWAS itself, which limits the power of the bipolar-conditional component to detect unique pleiotropic associations beyond those already captured by the shared factor or the schizophrenia-conditional component. Second, the independent recovery of 76 of the 99 novel pleiotropic loci in a substantially larger education GWAS^39^ constitutes an external replication that corroborates the validity of the PLEIO framework: these loci capture true polygenic signal at the intersection of psychiatric liability and cognitive phenotypes rather than inflated discovery through analytic artefact, and their detection here demonstrates that leveraging pleiotropic structure across genetically correlated traits can achieve comparable discovery power to univariate analyses conducted at considerably larger sample sizes.

Following the pleiotropic meta-analysis, we partitioned each component into concordant and discordant locus sets based on the direction of genetic correlations with CTP and EDU, yielding six locus sets: for SCZcondBIP, 439 concordant and 284 discordant loci; for BIPcondSCZ, 382 concordant and 268 discordant loci; and for the PSY-shared factor, 433 concordant and 487 discordant loci. Merging these six sets at a minimum overlap of one base pair yielded 867 consensus loci. The relative sizes of these six sets are themselves informative. Both SCZcondBIP and BIPcondSCZ are concordant-skewed (roughly 60:40 concordant:discordant), meaning that once shared psychosis liability is removed, the majority of disorder-specific loci align with the sign of that disorder’s overall genetic correlation with cognition (negative for SCZcondBIP, positive for BIPcondSCZ). By contrast, the PSY-shared factor is essentially balanced (433 concordant to 487 discordant), with discordant loci marginally more numerous. This balance is directly consistent with the mixed correlation profile of the shared factor (negative with CTP, positive with EDU), where neither cognitive direction predominates at the locus level. These distributional differences across components motivate the pathway-level dissection that follows.

Functional annotation of each of the six partitions revealed divergent biological architectures underlying cognitive trajectories across the disorders. Within SCZcondBIP, concordant loci (increased cognition and education, reduced schizophrenia risk) were enriched for neurodevelopmental and transcriptional pathways (sequence-specific DNA binding, positive regulation of RNA metabolism, GABAergic neuron differentiation, β-catenin binding, diencephalon and forebrain neuron differentiation, and cell-fate commitment) with a nominal trend toward early prenatal expression. This roots the reduced cognitive axis of schizophrenia-specific liability in early neurodevelopmental biology. It also provides a pathway-level substrate for the well-established observation that cognitive deficits in schizophrenia correlate with disruption of cortical-hippocampal maturation and inhibitory differentiation^40,41^. It is further supported by our previous finding that genes at loci linked to increased disease risk and reduced cognition are largely neurodevelopmental^28,29^. By contrast, SCZcondBIP discordant loci (increased cognition, increased schizophrenia risk) were enriched for cellular-homeostasis and metabolic-stress pathways (response to oxygen-glucose deprivation, nuclear protein-containing complex, UMP/dUMP metabolism) with no developmental enrichment. This branch of schizophrenia-specific risk may therefore operate outside classical neurodevelopmental windows, acting through nucleotide metabolism and stress resilience to influence synaptic plasticity. In our earlier work, schizophrenia discordant loci carried the synaptic-signalling signal^28,29^; here that signal instead appears in the shared discordant partition, suggesting synaptic-signaling pathways responsible for higher cognitive function and disease risk are probably driven by shared psychosis liability rather than by schizophrenia-specific discordant loci. The schizophrenia-cognition relationship therefore reflects at least two separable subprogrammes: an early developmental-regulatory branch (cognitive disadvantage, increased risk) and a later homeostatic/metabolic-stress branch involved in synaptic plasticity (cognitive advantage despite increased risk). In that sense, the latter may represent a pathway-level analogue of the semi-neurodegenerative or homeostatic vulnerability^42^ that clinically distinguishes schizophrenia from bipolar disorder.

BIPcondSCZ showed the mirror-image pattern. Concordant loci (increased cognition & education and increased bipolar liability) were enriched for synaptic-structure and signalling pathways (synaptic membrane adhesion, neurotrophin TRK receptor binding, cranial ganglion development), pointing to a biology of synaptic organization and connectivity rather than the transcriptional pathways observed in SCZcondBIP concordant loci. This is consistent with evidence that bipolar disorder, unlike schizophrenia, is not uniformly marked by premorbid cognitive impairment and can be associated with preserved or supranormal premorbid performance. Discordant loci (decreased cognition/education, increased bipolar liability) were enriched for pathways responsible for cellular homeostasis in response to stress (mitochondrial fragmentation in apoptosis, mitochondrial protein catabolism, and intrinsic apoptotic signaling in response to hypoxia). This pattern is notable because mitochondrial dynamics are integral to synaptic maintenance and neuronal bioenergetics. Mitochondrial dysfunction and reduced cellular resilience have been repeatedly implicated in bipolar pathophysiology and linked to synaptic dysfunction and progressive cognitive decline^43–46^. Genes associated with both concordant and discordant loci were robustly enriched across the early-, early-mid-, and late-mid-prenatal periods, indicating that bipolar-specific liability is fundamentally prenatal in developmental origin regardless of its cognitive direction. For the PSY-shared factor, concordant loci (increased shared risk, decreased cognition) implicated cellular-homeostasis and synaptic-plasticity pathways. Although these enrichments did not survive FDR correction, one of the strongest nominal signals was regulation of synaptic structure plasticity. This is directionally consistent with the disorder-specific reduced-cognition partitions, although the specific enrichments differ; SCZcondBIP toward neurodevelopmental pathways and BIPcondSCZ toward cellular homeostasis in response to stress. It also fits the broader psychosis literature, which states that cognitive impairment is central to psychotic illness and a major driver of functional outcome, but is marked by considerable inter-individual heterogeneity from the earliest stages of illness^47,48^. Discordant loci (increased cognition/education and increased shared psychosis liability) showed no FDR-significant pathways, but leading nominal signals (B-1 B-cell differentiation, B-cell proliferation in immune response, and synaptic vesicle loading) suggest a distinct branch in which cognitive advantage coexists with illness risk through neuroimmune and vesicular mechanisms.

The biological differentiation observed across partitions at the pathway level was further reflected in the drug-target enrichment analysis. At the nominal P value, each partition showed distinct pharmacological class biases: the SCZcondBIP concordant partition was enriched for antipsychotics and L-type calcium channel blockers, the SCZcondBIP discordant partition for anticonvulsant mood stabilisers, the BIPcondSCZ discordant partition for a broad mix of antipsychotics, dopaminergic agents, glutamate modulators, and anti-inflammatory compounds, and the PSY-shared discordant partition for immune and anti-inflammatory agents (celecoxib, curcumin, sulfasalazine, lenalidomide). Among cognitive-adjunct compounds, methylphenidate (enriched in three partitions, strongest at P = 0.003 in BIPcondSCZ discordant), buspirone (SCZcondBIP concordant), and pyridostigmine (PSY-shared concordant) reached P < 0.05.

Several of the enriched compounds have independent clinical support, including methylphenidate, buspirone, celecoxib and curcumin, and raloxifene. Broadly, pro-attentional/serotonergic agents clustered with partitions associated with increased cognition, anti-inflammatory and SERM/neurosteroid agents with the shared-psychosis axis, and anticonvulsant mood stabilisers with the high-cognition/high-SCZ-risk partition, mirroring the developmental-versus-homeostatic split observed in the pathway analyses. These observations are however exploratory and require cautious interpretation given the nominal significance threshold and the indirect nature of gene-to-drug inference in this framework.

## Conclusion

The genetic relationship between psychiatric liability and cognition is not unitary. Integrating disease-specific and shared psychosis components with endophenotypes such as cognitive and educational traits through directional pleiotropic meta-analysis reveals that clinical cognitive heterogeneity within and across schizophrenia and bipolar disorder reflects the relative contribution of dissociable biological branches rather than variation along a single liability dimension.

## Future directions

Partition-specific polygenic scores could be evaluated in deeply phenotyped clinical cohorts to test whether they stratify individuals by cognitive trajectory and functional outcome more effectively than DSM tool. Integrating the six partitions with cell-type-resolved and developmental-stage-resolved transcriptomic resources would further refine the biological splits identified here. Finally, applying the same directional pleiotropic decomposition to other intermediate phenotypes, including immune and metabolic traits, would test whether the substructure recovered here is specific to the cognitive axis or a general property of pleiotropic psychiatric risk.

## Limitations

The present study has several limitations. The analyses rely on European-ancestry GWAS, limiting generalisability. The drug-target enrichments were largely nominal and should be interpreted cautiously.

## Supporting information

supplementary table description

## Data Availability

All data produced in the present study are available upon reasonable request to the authors

