## supplementary table description for "Cognitive pleiotropy reveals disorder-specific and shared biology for schizophrenia and bipolar disorder"

[1]Northwell, New Hyde Park, NY, USA.

[2]Institute of Behavioral Science, Feinstein Institutes for Medical Research, Manhasset, NY, USA.

[3]Institute of Mental Health, Singapore

[4]Lee Kong Chian School of Medicine, Population and Global Health, Nanyang Technological University

[5]Departments of Psychiatry and Molecular Medicine, Zucker School of Medicine at Hofstra/Northwell, Hempstead, NY

*These authors contributed equally

§Corresponding Author:

Todd Lencz; phone: +1 917-453-4647;; Office: Division of Psychiatry Research, Zucker Hillside Hospital, Glen Oaks, NY, USA 11004

**Supplementary Table Description:**

**Supplementary Table 1a | Unpartitioned pleiotropic consensus loci.**

Independent genome-wide significant consensus loci identified after PLEIO meta-analysis of each component (SCZcondBIP, BIPcondSCZ, PSY-shared) jointly with cognitive task performance (CTP) and educational attainment (EDU). Columns report the consensus locus coordinates, binary component membership indicators, lead SNPs, and constituent member loci.

**Supplementary Table 1b | Concordant and discordant partitioned consensus loci.**

Consensus loci from the PLEIO meta-analysis partitioned into concordant (effect direction on the component aligns with the expected direction on CTP/EDU) and discordant (opposite direction) sets. Columns report the consensus locus identifier (ConsensusLocus), chromosome (chr), base-pair coordinates (start, end), locus number (locus), number of contributing partitions (nSets), binary presence indicators for each of the six partitions (SCZcondBIP concordant/discordant, BIPcondSCZ concordant/discordant, PSY-shared concordant/discordant), partition-specific lead SNPs (LeadSNP), and constituent member loci (members).

**Supplementary Table 2 | Novel pleiotropic loci.**

Assessment of novelty for loci identified after PLEIO meta-analysis of each component with CTP and EDU. A locus was designated as novel only if it did not overlap (≥1 bp) with genome-wide significant loci from any of the four input GWAS (schizophrenia, bipolar disorder, CTP, educational attainment EA3) and did not fall within 20 kb of a lead variant from the EA4 GWAS. Columns report the genetic component (Component), locus details (Locus, chr, start, end, region), lead SNP (LeadSNP), P-value (p), overlap indicators for each input study (in_SCZ, in_BIP, in_CTP, in_EA3), proximity to EA4 lead SNPs (near_EA4_leadSNP_20kb, EA4_leadSNP), and final novelty designation (Novelty).

**Supplementary Table 3 | MAGMA gene-set enrichment results.**

MAGMA (v1.10) competitive gene-set enrichment results for each of the six concordant/discordant partitions (SCZcondBIP concordant/discordant, BIPcondSCZ concordant/discordant, PSY-shared concordant/discordant). Columns report the gene-set name (FULL_NAME), followed by partition-specific metrics: number of mapped genes (_NGENES), enrichment effect size (_BETA), standardised effect size (_BETA_STD), standard error (_SE), nominal P-value (_P), and false discovery rate (_FDR).

**Supplementary Table 4 | BrainSpan developmental-timing enrichment.**
MAGMA gene-property analysis testing for developmental-stage-specific expression enrichment of FUMA-mapped genes across BrainSpan transcriptomic samples spanning early prenatal to late adult stages. Columns report the developmental period (Developmental period), followed by component- and partition-specific metrics: number of genes (NGENES), beta estimates (BETA, BETA_STD), standard errors (SE), nominal P-values (P), and FDR-adjusted P-values (FDR-P).

**Supplementary Table 5 | Enrichr drug-target enrichment.**
Enrichment of genes mapped to each component and partition against the Drug Gene Interaction Database (DGIdb) via Enrichr. Columns report the targeted term (Term), followed by partition-specific metrics: overlap count (Overlap), P-value (P-value), adjusted P-value (Adjusted P-value), historical P-values (Old P-value, Old Adjusted P-value), odds ratio (Odds Ratio), combined score (Combined Score), and implicated genes (Genes).

**Supplementary Tables 6a–f | DGIdb gene–drug interaction catalogues by partition.**
Drug–gene interactions for FUMA-mapped genes queried against DGIdb 5.0, reported separately for each partition (6a, SCZcondBIP concordant; 6b, SCZcondBIP discordant; 6c, BIPcondSCZ concordant; 6d, BIPcondSCZ discordant; 6e, PSY-shared concordant; 6f, PSY-shared discordant). Columns report the gene symbol (gene), drug name (drug), regulatory approval status (regulatory approval), clinical indication (indication), and interaction score (interaction score).
