## Supplementary Method for "Cognitive pleiotropy reveals disorder-specific and shared biology for schizophrenia and bipolar disorder"

[1]Northwell, New Hyde Park, NY, USA.

[2]Institute of Behavioral Science, Feinstein Institutes for Medical Research, Manhasset, NY, USA.

[3]Institute of Mental Health, Singapore

[4]Lee Kong Chian School of Medicine, Population and Global Health, Nanyang Technological University

[5]Departments of Psychiatry and Molecular Medicine, Zucker School of Medicine at Hofstra/Northwell, Hempstead, NY

*These authors contributed equally

§Corresponding Author:

Todd Lencz; phone: +1 917-453-4647;; Office: Division of Psychiatry Research, Zucker Hillside Hospital, Glen Oaks, NY, USA 11004

**Supplementary Methods:**

**GWAS summary statistics**

Building on the decomposition of schizophrenia and bipolar disorder into disease-specific (SCZcondBIP, BIPcondSCZ) and shared (PSY-shared) genetic components (Bhattacharyya et al., 2025), this study applies directionally informative pleiotropic meta-analysis to integrate each component with cognitive task performance and educational attainment. Briefly, SCZcondBIP and BIPcondSCZ were obtained by applying bidirectional multi-trait conditional and joint analysis (mtCOJO)[1] to genome-wide summary statistics from the Psychiatric Genomics Consortium (PGC) schizophrenia GWAS (76,755 cases and 243,649 controls, European ancestry)[2] and bipolar disorder GWAS (57,607 cases and 371,549 controls, European ancestry)[3]. mtCOJO conditions the effect-size estimates of one trait on the genetic effects of a second trait using GWAS summary statistics and an LD reference panel, thereby removing the shared genetic signal between the two disorders and isolating disorder-specific genetic architecture. PSY-shared was derived by fitting a single common-factor model to the schizophrenia and bipolar disorder GWAS within the Genomic Structural Equation Modeling (Genomic SEM)[4] framework, and extracting the common factor representing the latent genetic architecture shared between the two disorders. Full derivation, model-fit statistics, and validation of these three components, including their divergent patterns of genetic correlation with cognitive, cardiometabolic, and inflammatory phenotypes, are reported in our previous paper (MEDRXIV/2026/357436).

For the cognitive phenotypes, we obtained publicly available GWAS summary statistics for cognitive task performance (CTP; N = 260,576 individuals of European ancestry, meta-analysis of 57 cohorts assessing general cognitive ability using diverse neuropsychological instruments)[5] and educational attainment measured as years of schooling completed (EDU; N = 766,345 individuals of European ancestry)[6]. These two phenotypes were selected because they are genetically correlated yet partially non-overlapping in their genetic architecture ($r_g \approx 0.72$)[7], and because the three genetic components show markedly divergent correlation profiles with CTP and EDU: SCZcondBIP is negatively correlated with both, BIPcondSCZ is positively correlated with both, and PSY-shared retains the mixed profile seen at the disorder level (negative with CTP, positive with EDU). This divergence means that CTP and EDU jointly index distinct genetic influences on each component, making their simultaneous integration in a pleiotropic framework well suited to resolve the directional heterogeneity underlying cognitive outcomes across the two disorders, as demonstrated in our prior work applying this approach to schizophrenia[7,8].

### Quality control and harmonization

Each set of GWAS summary statistics was subjected to a standardized quality-control pipeline prior to analysis. Summary statistics were harmonized using the gwas2vcf tool[9,10], which converts GWAS results to Variant Call Format (VCF), standardizes allele representation to the GRCh37 reference genome, and ensures a consistent direction of reported SNP effects. During harmonization, we verified that all variants had non-zero effect-size estimates (beta ≠ 0) and non-zero standard errors (SE > 0); variants failing either criterion were removed.

The following variant-level filters were then applied sequentially: (i) insertion-deletion polymorphisms (indels) were removed, as these are less reliably imputed and mapped across studies; (ii) strand-ambiguous single-nucleotide polymorphisms (A/T and C/G SNPs) were removed to prevent allele-alignment errors; (iii) variants with imputation quality score (INFO) ≤ 0.3 were removed to exclude poorly imputed variants; and (iv) variants with minor allele frequency (MAF) ≤ 0.01 were removed, as low-frequency variants have inflated standard errors in GWAS of this sample size range and can produce spurious pleiotropic signals. The extended major histocompatibility complex (MHC) region on chromosome 6 (positions 25,000,000–35,000,000 bp, GRCh37) was excluded from all analyses because its exceptionally complex and extensive linkage-disequilibrium structure violates the assumptions of LD-based methods used throughout this study, including LD Score Regression, PLEIO, and MAGMA. After quality control, the intersection of variants present across all five input datasets (SCZcondBIP, BIPcondSCZ, PSY-shared, CTP, and EDU) was retained for analysis.

The 1000 Genomes Project Phase 3 European-ancestry panel (503 individuals, GRCh37) served as the linkage-disequilibrium reference for all analyses requiring LD estimation, including LD Score Regression preprocessing, PLEIO meta-analysis, FUMA locus definition, and MAGMA gene-based tests.

### Pleiotropic meta-analysis using PLEIO

Directionally informative pleiotropic meta-analyses were performed using the Pleiotropic Locus Exploration and Interpretation using Optimal test (PLEIO)[11]. Unlike conventional fixed-effect meta-analysis, which assumes that a variant affects all traits in the same direction and with proportional magnitude, PLEIO employs a variance-component framework that models heterogeneity in effect sizes and directions across traits, maximizing power to detect pleiotropic loci regardless of whether their effects are concordant or discordant across the input phenotypes. The PLEIO test statistic at each variant aggregates evidence for association across all input traits while accounting for the genetic and environmental correlation structure among those traits, yielding a single omnibus P value for pleiotropic association.

Three meta-analyses were conducted in parallel, each combining one genetic liability component with both cognitive phenotypes:

1. SCZcondBIP–CTP–EDU: the schizophrenia-conditional component jointly analyzed with cognitive task performance and educational attainment.

2. BIPcondSCZ–CTP–EDU: the bipolar-conditional component jointly analyzed with cognitive task performance and educational attainment.

3. PSY-shared–CTP–EDU: the shared psychosis factor jointly analyzed with cognitive task performance and educational attainment.

For each meta-analysis, the inter-trait genetic covariance matrix and environmental (residual) correlation matrix were estimated using the ldsc_preprocess.py script distributed with the PLEIO software package[11], which internally calls LD Score Regression (LDSC)[12] to estimate genetic correlations and heritabilities from the input GWAS summary statistics. These matrices are required by PLEIO to correctly model the non-independence of test statistics arising from phenotypic and genetic overlap among the input traits, thereby controlling type I error.

Genome-wide significance was defined at $P < 5 \times 10^{-8}$ for the omnibus PLEIO test statistic.

**Estimation of trait-specific effect directions using BLUP**

For each of the three PLEIO meta-analyses, per-variant trait-specific effect estimates were obtained using the best linear unbiased predictor (BLUP) functionality implemented within PLEIO (invoked via the --blup flag). The BLUP procedure leverages the estimated inter-trait genetic covariance structure to produce shrinkage-corrected, per-variant effect-size estimates for each input trait, conditional on the observed association evidence across all traits simultaneously. These BLUP estimates provide the direction and magnitude of each variant's contribution to each individual trait (i.e., to the liability component, to CTP, and to EDU), enabling the subsequent classification of pleiotropic variants into concordant and discordant categories based on their directional effect profile.

The BLUP approach is preferable to using raw marginal GWAS effect estimates for directional classification because it accounts for the correlation structure among traits and thereby reduces noise-driven misclassification of effect directions, particularly for variants with modest individual-trait associations that nonetheless contribute to the pleiotropic signal. This approach was previously validated in our work partitioning schizophrenia risk loci[7,8].

**Classification of pleiotropic loci into concordant and discordant sets**

The three liability components exhibit distinct patterns of genetic correlation with the two cognitive phenotypes: SCZcondBIP is negatively correlated with both CTP and EDU; BIPcondSCZ is positively correlated with both; and PSY-shared is negatively correlated with CTP but positively correlated with EDU. Because these correlation structures differ, the definition of "expected" versus "unexpected" allelic configurations must be specified separately for each component. The classification procedure, applied at the variant level using the BLUP effect estimates described above, was as follows.

SCZcondBIP (negative correlation with both CTP and EDU): Given the negative genetic correlation between SCZcondBIP and both cognitive traits, the biologically expected (concordant) configuration is one in which alleles that improve cognitive performance and educational attainment simultaneously reduce schizophrenia-specific liability. To facilitate interpretation under this framework, the sign of the BLUP effect estimate for SCZcondBIP was reversed prior to classification, as in our previous work[7,8]. After sign reversal, concordant variants were defined as those for which the BLUP effects on CTP, EDU, and (sign-reversed) SCZcondBIP were all positive—that is, variants where alleles that increase cognitive performance and education also decrease SCZcondBIP liability. Discordant variants were defined as those for which BLUP effects on CTP and EDU were positive but the (sign-reversed) SCZcondBIP BLUP effect was negative—that is, variants where alleles that increase cognitive performance and education paradoxically also increase schizophrenia-specific liability.

BIPcondSCZ (positive correlation with both CTP and EDU): Given the positive genetic correlation between BIPcondSCZ and both cognitive traits, no sign reversal was applied. Concordant variants were defined as those for which BLUP effects on CTP, EDU, and BIPcondSCZ were all positive—that is, variants where alleles that increase cognitive performance and education also increase bipolar-specific liability, consistent with the positive genetic correlation. Discordant variants were defined as those for which BLUP effects on CTP, EDU, and BIPcondSCZ were all negative—that is, variants where alleles that decrease cognitive performance and education also decrease bipolar-specific liability. Although the direction of co-movement (both decreasing) is consistent with a positive correlation in algebraic terms, these variants are classified as discordant because they represent the cognitive-disadvantage pole of bipolar-specific liability, opposing the clinically observed association between bipolar disorder and preserved or enhanced premorbid cognitive functioning.

PSY-shared (negative correlation with CTP, positive correlation with EDU): The mixed correlation profile of PSY-shared with respect to the two cognitive phenotypes required careful handling. Because PSY-shared resembles raw schizophrenia in its correlation structure (negative with CTP, positive with EDU), the sign of the BLUP effect estimate for PSY-shared was reversed prior to classification, analogous to the procedure for SCZcondBIP. And our previous work[7,8] After sign reversal, concordant variants were defined as those for which BLUP effects indicated that alleles decrease CTP and/or increase EDU while also increasing PSY-shared liability (i.e., the expected direction given the mixed correlation). Discordant variants were defined as those for which alleles increase CTP and/or decrease EDU while also increasing PSY-shared liability, representing the unexpected direction.

Null-effect threshold: Across all three classifications, per-trait BLUP effect estimates with absolute values smaller than 0.001 were treated as null (effectively zero), ensuring that only variants with directionally meaningful contributions to each trait were assigned to concordant or discordant categories. Variants for which any trait-specific BLUP estimate fell below this threshold were excluded from directional classification but remained in the overall PLEIO results.

**Genomic risk-locus definition and candidate-gene mapping (FUMA)**

Genome-wide significant variants ($P < 5 \times 10^{-8}$) from each of the three PLEIO meta-analyses, and from each of the six concordant/discordant partitions, were processed through the Functional Mapping and Annotation of Genetic Associations (FUMA) platform, version 2.1.5[13], to define genomic risk loci and identify candidate genes.

**Risk-locus definition:** FUMA defines risk loci through a two-stage clumping procedure. First, independent significant SNPs were identified as variants reaching $P < 5 \times 10^{-8}$ that are independent of each other at a linkage-disequilibrium threshold of $r^2 < 0.6$. Second, among independent significant SNPs, lead SNPs were identified by further clumping at $r^2 < 0.1$. Genomic risk loci were then defined by merging lead SNPs (and their associated LD-defined regions) that were located within 250 kb of each other into single loci. The LD reference panel for clumping was the 1000 Genomes Project Phase 3 European-ancestry panel. The boundaries of each risk locus were defined by the most extreme positions of all candidate SNPs (those in LD with independent significant SNPs at $r^2 \geq 0.6$ and with $P < 0.05$) within each locus.

**Candidate-gene mapping:** Within each defined risk locus, candidate genes were prioritized using three complementary mapping strategies implemented in FUMA: (i) Positional mapping: genes were assigned to a risk locus if any part of the gene body fell within a 10-kb window of the locus boundaries. This captures genes in immediate genomic proximity to the association signal. (ii) Expression quantitative trait locus (eQTL) mapping: SNPs within risk loci were linked to genes whose expression they significantly regulate (at FDR < 0.05) across multiple tissue panels. eQTL data sources included all data sources available in FUMA version 2.1.5. A SNP–gene pair was included if the SNP was a significant eQTL for that gene in at least one tissue. (iii) Three-dimensional chromatin-interaction mapping: SNPs were linked to genes through significant chromatin contacts identified in Hi-C datasets, including data from all sources available in FUMA version 2.1.5. A chromatin interaction was considered significant if it connected a genomic region containing a candidate SNP to a region overlapping a gene promoter (defined as 250 bp upstream and 500 bp downstream of the transcription start site), with FDR $< 1 \times 10^{-6}$. The union of genes identified across the three mapping strategies constituted the final candidate gene set for each PLEIO meta-analysis and for each concordant/discordant partition. These gene sets were carried forward to all downstream functional analyses.

**Locus overlap analysis and identification of novel pleiotropic loci**

To quantify the degree of locus sharing across the three liability components and to identify pleiotropic loci not previously reported in individual disorder or cognitive GWAS, we performed systematic locus-overlap analyses using BEDTools (version 2.30.0)[14].

**Cross-component overlap:** The genome-wide significant risk loci defined by FUMA for each of the three PLEIO meta-analyses were compared pairwise and jointly. Overlap between two loci was defined as a minimum of one shared base position (1-bp overlap using BEDTools intersect). Loci were classified into the following categories: (i) consensus loci, defined as the non-redundant union of all risk loci across the three PLEIO meta-analyses (obtained using BEDTools merge); (ii) loci shared across all three components (present in SCZcondBIP, BIPcondSCZ, and PSY-shared PLEIO results); (iii) loci shared between exactly two components; and (iv) loci unique to a single component.

The same overlap procedure was applied to the six directional partitions (concordant and discordant for each component) to assess convergence among same-direction locus sets (e.g., concordant across all three components) and divergence between opposite-direction sets.

**Novel locus identification:** To identify pleiotropic loci that emerge only through the joint analysis of liability components with cognitive phenotypes, the consensus loci from the three PLEIO meta-analyses were intersected with the published genome-wide significant loci from each of the following GWAS:

- Schizophrenia (PGC3)[2]

- Bipolar disorder (PGC-BIP)[3]

- Cognitive task performance[5]

- Educational attainment, third wave (EA3)[6]

- Educational attainment, fourth wave (EA4)[15]

Where a published GWAS provided summary statistics without pre-defined genomic risk-locus boundaries, loci were first delineated by processing the summary statistics through FUMA under default settings (as described above) to obtain comparable locus definitions.

A PLEIO locus was classified as novel if it satisfied all of the following criteria: (i) it did not overlap (at ≥1-bp resolution) any genome-wide significant locus from the schizophrenia, bipolar disorder, cognitive task performance, or EA3 GWAS; and (ii) it did not lie within 20 kb of any genome-wide significant lead variant reported in EA4. The EA4 GWAS was included as a comparison despite not being used as input to the PLEIO meta-analyses because it represents the most recent and best-powered educational-attainment GWAS available (N = 3,037,499), and novelty claims require exclusion of loci discoverable by any of the constituent phenotypes at maximal statistical power. The 20-kb buffer for EA4 lead SNP was applied because full summary satistics were not available for EA4[15].

**Competitive gene-set enrichment analysis (MAGMA)**

Gene-set enrichment analysis was performed using Multi-marker Analysis of GenoMic Annotation (MAGMA, version 1.10)[16] to identify biological pathways and functional categories over-represented among genes implicated by each liability component and its directional partitions.

**Gene annotation:** SNPs were annotated to 18,877 protein-coding genes (NCBI build 37, gene-boundary definitions from the NCBI gene database) using an asymmetric window extending 35 kb upstream and 10 kb downstream of each gene body[17]. The upstream extension was chosen to capture promoter regions and proximal enhancers, while the downstream extension captures 3' regulatory elements.

**Gene-based association testing:** For each of the six concordant/discordant partitions (concordant and discordant for each of SCZcondBIP, BIPcondSCZ, and PSY-shared), gene-level association statistics were computed using the MAGMA SNP-wise mean model, which combines the association P values of all SNPs annotated to a gene into a single gene-level test statistic. Inter-SNP linkage disequilibrium was modeled using the 1000 Genomes Project Phase 3 European panel to ensure that correlated SNPs within a gene do not inflate the gene-level statistic.

Competitive gene-set testing: Prior to competitive testing, genes located on the sex chromosomes (X and Y) and within the extended MHC region (chromosome 6: 25–35 Mb) were removed from all gene sets to prevent confounding by sex-linked effects and the complex LD structure of the MHC, respectively. Gene sets containing fewer than five remaining genes after this filtering were excluded to ensure stable enrichment estimates. The filtered gene-level statistics were then tested for competitive enrichment against curated gene sets from the Gene Ontology (GO) collection of the Molecular Signatures Database (MSigDB, version 2024.1) C5 collection[18], encompassing three ontology domains: Biological Process (BP), Cellular Component (CC), and Molecular Function (MF). Competitive testing evaluates whether genes within a given set show stronger associations with the phenotype than genes outside the set, after accounting for gene size, gene density, and other potential confounders—as opposed to self-contained testing, which evaluates only whether genes in the set are associated at all.

Multiple-testing correction was applied using the Benjamini–Hochberg procedure to control the false discovery rate (FDR) at 5% across all tested gene sets within each of the six analyses.

**Drug-target enrichment analysis**

To identify whether genes implicated by each concordant and discordant partition are disproportionately targeted by existing pharmacological agents, the FUMA-prioritized candidate genes for each of the six gene sets were submitted to the Enrichr platform[19–21] for enrichment testing against the Drug–Gene Interaction Database (DGIdb)[22]. Enrichr computes enrichment using a Fisher exact test, comparing the overlap between the input gene set and each drug-target gene set in DGIdb against the background of all human protein-coding genes, with results ranked by adjusted P value. This analysis identifies drug categories and individual compounds whose molecular targets are over-represented among the implicated genes, providing an initial assessment of pharmacological relevance and potential for drug repurposing.

**Druggability assessment**

To provide a more granular assessment of the therapeutic tractability of genes implicated by each directional partition, the FUMA-mapped candidate genes from all six concordant/discordant gene sets were individually annotated against the Drug–Gene Interaction module of DGIdb (version 5.0)[22]. For each gene, we retrieved: (i) all known drug–gene interactions; (ii) the regulatory-approval status of each interacting drug (e.g., FDA-approved, EMA-approved, investigational, or withdrawn); (iii) the primary clinical indication for each approved drug; and (iv) the interaction type (e.g., inhibitor, agonist, modulator, antibody). A gene was classified as "druggable" if it was the annotated molecular target of at least one drug with current regulatory approval by the FDA or EMA for any clinical indication.

For each of the six gene sets, we computed the following summary metrics: (i) the total number of unique drug–gene interactions; (ii) the number and proportion of genes classified as druggable (targeted by ≥1 approved drug); (iii) the distribution of interacting drugs across therapeutic categories (e.g., antipsychotics, mood stabilizers, antiepileptics, immunomodulators, oncology agents); and (iv) the interaction types represented. These metrics were compared across the six groups (concordant and discordant for each of the three components). Because gene–drug interaction counts in DGIdb are influenced by gene-set size, by the degree of prior biological characterization of constituent genes, and by historical research emphasis on certain gene families (e.g., ion channels, kinases, G-protein-coupled receptors), formal statistical comparisons of druggability proportions across groups were not performed. Instead, differences in druggability and drug-class composition are reported descriptively to highlight potential translational opportunities and to identify whether concordant and discordant gene sets within each component implicate pharmacologically distinct target classes.


1. Zhu, Z. *et al.* Causal associations between risk factors and common diseases inferred from GWAS summary data. *Nat. Commun.* **9**, 224 (2018).
2. Trubetskoy, V. *et al.* Mapping genomic loci implicates genes and synaptic biology in schizophrenia. *Nature* **604**, 502–508 (2022).
3. O'Connell, K. S. *et al.* Genomics yields biological and phenotypic insights into bipolar disorder. *Nature* **639**, 968–975 (2025).
4. Grotzinger, A. D. *et al.* Genomic structural equation modelling provides insights into the

multivariate genetic architecture of complex traits. *Nat. Hum. Behav.* **3**, 513–525 (2019).

5. Lam, M. *et al.* Identifying nootropic drug targets via large-scale cognitive GWAS and transcriptomics. *Neuropsychopharmacology* **46**, 1788–1801 (2021).
6. Lee, J. J. *et al.* Gene discovery and polygenic prediction from a genome-wide association study of educational attainment in 1.1 million individuals. *Nat. Genet.* **50**, 1112–1121 (2018).
7. Lam, M. *et al.* Pleiotropic meta-analysis of cognition, education, and schizophrenia differentiates roles of early neurodevelopmental and adult synaptic pathways. *Am. J. Hum. Genet.* **105**, 334–350 (2019).
8. Bhattacharyya, U., John, J., Lencz, T. & Lam, M. Dissecting schizophrenia biology using pleiotropy with cognitive genomics. *Biol. Psychiatry* **98**, 670–678 (2025).
9. Lyon, M. S. *et al.* The variant call format provides efficient and robust storage of GWAS summary statistics. *Genome Biol.* **22**, 32 (2021).
10. Elsworth, B. *et al.* The MRC IEU OpenGWAS data infrastructure. *bioRxiv* (2020) doi:10.1101/2020.08.10.244293.
11. Lee, C. H., Shi, H., Pasaniuc, B., Eskin, E. & Han, B. PLEIO: a method to map and interpret pleiotropic loci with GWAS summary statistics. *Am. J. Hum. Genet.* **108**, 36–48 (2021).
12. Bulik-Sullivan, B. K. *et al.* LD Score regression distinguishes confounding from polygenicity in genome-wide association studies. *Nat. Genet.* **47**, 291–295 (2015).
13. Watanabe, K., Taskesen, E., van Bochoven, A. & Posthuma, D. Functional mapping and annotation of genetic associations with FUMA. *Nat. Commun.* **8**, 1826 (2017).
14. Quinlan, A. R. & Hall, I. M. BEDTools: a flexible suite of utilities for comparing genomic features. *Bioinformatics* **26**, 841–842 (2010).
15. Okbay, A. *et al.* Polygenic prediction of educational attainment within and between families from genome-wide association analyses in 3 million individuals. *Nat. Genet.* **54**, 437–449

(2022).

16. de Leeuw, C. A., Mooij, J. M., Heskes, T. & Posthuma, D. MAGMA: generalized gene-set analysis of GWAS data. *PLoS Comput. Biol.* **11**, e1004219 (2015).
17. Singh, T. *et al.* Rare coding variants in ten genes confer substantial risk for schizophrenia. *Nature* **604**, 509–516 (2022).
18. MSigDB. https://www.gsea-msigdb.org/gsea/msigdb/collections.jsp.
19. Xie, Z. *et al.* Gene set knowledge discovery with Enrichr. *Curr. Protoc.* **1**, e90 (2021).
20. Kuleshov, M. V. *et al.* Enrichr: a comprehensive gene set enrichment analysis web server 2016 update. *Nucleic Acids Res.* **44**, W90–7 (2016).
21. Chen, E. Y. *et al.* Enrichr: interactive and collaborative HTML5 gene list enrichment analysis tool. *BMC Bioinformatics* **14**, 128 (2013).
22. Cannon, M. *et al.* DGIdb 5.0: rebuilding the drug-gene interaction database for precision medicine and drug discovery platforms. *Nucleic Acids Res.* **52**, D1227–D1235 (2024).
